# Sex and gender differences in adverse events following receipt of influenza and COVID-19 vaccination among healthcare workers

**DOI:** 10.1101/2024.01.17.24301440

**Authors:** Anna Yin, Nadia Wang, Patrick J. Shea, Erica N. Rosser, Helen Kuo, Janna R. Shapiro, Katherine Z.J. Fenstermacher, Andrew Pekosz, Richard E. Rothman, Sabra L. Klein, Rosemary Morgan

**Author notes:** **Corresponding author contact** Sabra L. Klein and Rosemary Morgan.

## Abstract

**Introduction:** Active and passive surveillance studies have found that a greater proportion of females report adverse events (AE) following receipt of either the COVID-19 or seasonal influenza vaccine compared to males. We sought to determine the intersection of biological sex and sociocultural gender differences in prospective active reporting of vaccine outcomes, which remains poorly characterized.

**Methods:** This cohort study enrolled Johns Hopkins Health System healthcare workers (HCWs) who were recruited from the annual fall 2019-2022 influenza vaccine and the fall 2022 COVID-19 bivalent vaccine campaigns. Vaccine recipients were enrolled the day of vaccination and AE surveys were administered two days post-vaccination (DPV) for bivalent COVID-19 and Influenza vaccine recipients. Data were collected regarding the presence of a series of solicited local and systemic AEs. Open-ended answers about participants’ experiences with AEs also were collected for the COVID-19 vaccine recipients.

**Results:** Females were more likely to report local AEs after influenza (OR=2.28, p=0.001) or COVID-19 (OR=2.57, p=0.008) vaccination compared to males, regardless of age or race. Males and females had comparable probabilities of reporting systemic AEs after influenza (OR=1.18, p=0.552) or COVID-19 (OR=0.96, p=0.907) vaccination. Exogenous hormones from birth control use did not impact the rates of reported AEs following COVID-19 vaccination among reproductive-aged female HCWs. Women reported more interruptions in their daily routine following COVID-19 vaccination than men and were more likely to seek out self-treatment. More women than men scheduled their COVID-19 vaccination before their days off in anticipation of AEs.

**Conclusions:** Our findings highlight the need for sex- and gender-inclusive policies to inform more effective occupational health vaccination strategies. Further research is needed to evaluate the potential disruption of AEs on occupational responsibilities following mandated vaccination for healthcare workers and to more fully characterize the post-vaccination behavioral differences between men and women.

**KEY MESSAGE:** *What is already known on this topic:*

⇒ Among diversely aged adults 18-64 years, females report more AEs to vaccines, including the influenza and COVID-19 vaccines, than males.
⇒ Vaccine AEs play a role in shaping vaccine hesitancy and uptake.
⇒ Vaccine uptake related to influenza and COVID-19 are higher among men than women.
⇒ Research that addresses both the sex and gender disparities of vaccine outcomes and behaviors is lacking.

*What this study adds:*

⇒ This prospective active reporting study uses both quantitative and qualitative survey data to examine sex and gender differences in AEs following influenza or COVID-19 vaccination among a cohort of reproductive-aged healthcare workers.

*How this study might affect research, practice, or policy:*

⇒ Sex and gender differences in AEs and perceptions relating to vaccination should drive the development of more equitable and effective vaccine strategies and policies in occupational health settings.

## INTRODUCTION

Females report more adverse events (AEs) than males to many vaccines, including the influenza[1-3] and COVID-19[1-5] vaccines. These differences have been attributed to biological differences between males and females (e.g., sex steroid effects on inflammatory immune responses) as well as gender differences (e.g., the socio-cultural differences between men and women), including gender reporting bias[2, 4-10], with few studies considering both sex and gender facets in the same study population[2]. AEs occur when the body mounts an immune response to the vaccine antigen, increasing secretion of inflammatory cytokines and recruitment of immune cells to the injection site, which can also enter the bloodstream and lead to more systemic AEs, such as fever, malaise, and fatigue[11]. Sex steroid hormones (e.g., estrogens, androgens, and progesterone) and their receptors have been hypothesized as critical regulators of immune cell responses that cause differential cytokine secretion between males and females[10, 12-15]. In response to infection and vaccination, females have been shown to have greater immune activation, higher production of antibodies, and increased T cell activation, possibly making them more likely to experience AEs compared to males[5, 10, 12, 14, 16, 17].

Beyond the physical impacts of AEs, experiences with AEs following vaccination can influence vaccine attitudes and patterns of uptake[18, 19]. The World Health Organization lists vaccine hesitancy as one of the top ten threats to global health[20]. Vaccine hesitancy related to influenza and COVID-19 is higher among women than men, which has been hypothesized to be due to the increased likelihood of AEs in females than males[21-25]. Men consistently have higher influenza vaccine acceptance than women, with White men often having less hesitancy than either Black or White women or Black men[26-28]. Similarly, male healthcare workers (HCWs) have also been found to have lower COVID-19 vaccine hesitancy with higher vaccine uptake than women[29-32].

In the United States, the Centers for Disease Control (CDC) recommends annual influenza vaccination for anyone aged 6 months or older[33]. Among HCWs, many employment or state laws require receipt of annual influenza vaccination to slow disease transmission between providers and patients[34]. During the COVID-19 pandemic, COVID-19 vaccination was nationally mandated for HCWs as terms of employment[35]. Despite SARS-CoV-2 becoming endemic with consistent spread and mutations noted[36, 37], COVID-19 vaccination requirements were terminated for HCWs when federal legislation lifted the public health emergency in May 2023[35]; those policy changes have and are anticipated to significantly impact future vaccine uptake. During the COVID-19 pandemic, we conducted a cohort study through the Johns Hopkins Health System to explore sex and gender differences in active, self-reported AEs following both seasonal quadrivalent influenza and bivalent COVID-19 vaccines. We further explored AEs by race/ethnicity and age among the influenza and COVID-19 vaccine recipients. Gender-related responses were collected with open-ended questions about AEs after receipt of the COVID-19 vaccine among women and men. Our goal was to provide a thorough assessment of sex and gender differences in AE reporting among HCWs to improve policies and messaging around mandatory vaccine programs.

## METHODS

### Study design and participants

Our study involved two separate survey-based cohorts of human participants, which were approved by the Johns Hopkins Institutional Review Boards (IRB00259171, IRB00091667). Consent was obtained for all participants as part of the enrollment process. Influenza vaccination has been a long-standing requirement at Johns Hopkins for anyone working directly with patients or in a clinical setting; additionally, during the pandemic from 2020-2022, a policy requiring the same workers to receive COVID-19 vaccination was established. Reproductive-aged adult (18-49) HCWs of the Johns Hopkins Health System (JHHS) receiving the inactivated quadrivalent influenza vaccine were considered eligible. Adult (≥18) HCWs receiving the 2022 Pfizer-BioNTech ancestral/Omicron BA.5 bivalent COVID-19 vaccine were also considered eligible. Influenza vaccine AE data was collected as part of a larger study designed to compare immunological vaccine responses by sex with equal sample sizes of females and males enrolled. HCWs were recruited by fliers, emails, and announcements about the annual vaccination program and were enrolled upon receipt of the influenza vaccine at the hospital. For the COVID-19 vaccine study, participants were recruited using flyers distributed at the time of vaccination at key locations around the hospital.

### Data collection

Annual influenza AE vaccination data were collected from September through October of 2019-2022. COVID-19 vaccine AE data was collected from September through October 2022. Influenza vaccine AE survey forms, provided as a hard copy at the time of consent/enrollment, were to be completed within two days post-vaccination (DPV) by participants and returned to study coordinators in-person at their next scheduled visit. COVID-19 bivalent vaccine AE surveys were electronically administered and collected via REDCap at two DPV for those who agreed to participate at the time of vaccination. Participants’ experience of local AEs at the site of injection (i.e., warmth, redness, swelling, short-duration pain, long-duration pain, and itchiness), and systemic AEs (i.e., sweating, malaise, muscle aches, insomnia, headaches, fever, and chills) were collected as yes/no answers and tallied by category. The level of inconvenience was measured by multiple choice answers. Open-ended questions were included within the COVID-19 vaccine study to explore reasons for vaccine uptake and responses to AEs. All survey responses and demographics were self-reported by participants. Male and female terminology was used to refer to biological differences. Man and woman terminology was used to refer to gender differences in behaviors or outcomes.

### Quantitative statistical analysis

Statistical analyses were performed using Stata 17.0 and GraphPad Prism. Any AE was defined as having at least one local or systemic AE. Sex differences in the reporting of AEs were analyzed using logistic regression models. Interaction terms were also included in the model to examine age, race/ethnicity, or hormonal birth control effects on the probabilities of AEs by sex. Probabilities were plotted along with 95% confidence intervals by sex. P-values less than 0.05 were considered statistically significant.

### Qualitative analysis of open-ended questions within COVID-19 vaccine AE survey

Open-ended questions regarding reasons for vaccine uptake, and response to AEs were analyzed using thematic analysis. Completed free-response answers from participants were explored and grouped into themes and responses from men and women were compared.

## RESULTS

### Participant characteristics

A total of 300 influenza vaccines (n=50 for females and n=50 for males per year) were administered across the three years (2019-20, 2021-22, and 2022-23) with AE data available for 265 (88%) of the participants and missing for 35 (12%; **Table 1**). Of these, 50.2% were female (n=133) and 49.8% male (n=132). The average age across the study years was 30.75 years. Participants were predominately White at 60.8% (n=160), followed by Asian at 19.4% (n=51), with 12.9% Black or African American (n=34). For the influenza cohort, 13.2% (n=35) identified as Hispanic or Latino.

**Table 1.** Study participant demographics.

| Influenza Vaccine Cohort |  |  |  |  | COVID-19 Vaccine Cohort |
| --- | --- | --- | --- | --- | --- |
| Season | 2019-20 | 2021-22 | 2022-23 | Total | 2022-23 |
| **Sex, n (%)** |  |  |  |  |  |
| Male | 45 (50.6%) | 40 (45.45%) | 47 (53.4%) | 132 (49.8%) | 46 (23.5%) |
| Female | 44 (49.4%) | 48 (54.55%) | 41 (46.6%) | 133 (50.2%) | 150 (76.5%) |
| **Ethnicity, n (%)** |  |  |  |  |  |
| Hispanic or Latino | 15 (16.9%) | 11 (12.5%) | 9 (10.2%) | 35 (13.2%) | n/a |
| **Race, n (%)** |  |  |  |  |  |
| White | 44 (50.6%) | 67 (76.1%) | 49 (55.7%) | 160 (60.8%) | 127 (64.8%) |
| Asian | 15 (17.2%) | 12 (13.6%) | 24 (27.3%) | 51 (19.4%) | 41 (20.9%) |
| Black | 16 (18.4%) | 6 (6.8%) | 12 (13.6%) | 34 (12.9%) | 17 (8.7%) |
| American Indian | 2 (2.3%) | 1 (1.1%) | 0 (0%) | 3 (1.1%) | 0 (0%) |
| Native Hawaiian | 0 (0%) | 2 (2.3%) | 0 (0%) | 2 (0.8%) | 0 (0%) |
| Other | 8 (9.2%) | 0 (0%) | 3 (3.4%) | 11 (4.2%) | 11 (5.6%) |
| Unknown | 2 (2.3%) | 0 (0%) | 0 (0%) | 2 (0.8%) | 0 (0%) |
| **Age, mean (SD)** | 30.45 (6.9) | 31.01 (6.7) | 30.8 (7.6) | 30.75 (7.0) | 38.4 (12.3) |
| **Any AE, n (%)** | 63 (71.8%) | 55 (62.5%) | 46 (52.3%) | 164 (61.9%) | 169 (86.2%) |
| **Any local AE, n (%)** | 52 (58.4%) | 41 (46.6%) | 36 (40.9%) | 129 (48.7%) | 138 (70.4%) |
| **Any systemic AE, n (%)** | 24 (27.0%) | 26 (29.6%) | 20 (22.7%) | 70 (26.4%) | 118 (60.2%) |
| **Total** | 89 (33.6%) | 88 (33.2%) | 88 (33.2%) | 265 | 196 |

For the COVID-19 bivalent vaccine cohort, 212 HCWs enrolled and received vaccination with AE survey data missing for 16 (8%) and available for 196 (92%) of those participants, consisting of 76.5% (n=150) females and 23.5% (n=46) males (**Table 1**). The average age was 38.4 years and the cohort predominately consisted of White participants at 64.8% (n=160) followed by Asian participants at 20.9% (n=41), and Black/African American participants at 8.7% (n=17).

### AEs were predominately localized and mild

Among the 265 total influenza HCW recipients across the three years, 164 (62%) reported having at least one AE with 57% (n=94) having only local AEs, 21% (n=35) having only systemic AEs, and 21% (n=35) having both local and systemic AEs. Of the 178 that responded to the question about level of inconvenience, the majority of recipients (n=142, 80%) did not experience any inconvenience when surveyed two DPV. Seventeen percent of HCWs (n=31) reported mild inconvenience where they were able to do 75-99% of their daily activities, 2% (n=4) reported moderate inconvenience where they were able to do 25-75% of their daily activities, and only 1% (n=1) reported severe inconvenience with capacity to do 0-25% of their daily activities.

Among the 196 COVID-19 bivalent vaccine recipients in 2022, 86% (n=169) of participants reported at least one AE. Of those, 30% reported only local AEs, 18% reported only systemic AEs, and 51% reported having both local and systemic AEs. The majority (53%, n=90) did not experience any inconvenience with their daily activities. Twenty-three percent reported mild inconvenience where they were able to do 75-99% of their daily activities. Eighteen percent of HCWs reported moderate inconvenience and were able to do 25-75% of their daily activities. Only 5% reported being severely inconvenienced with the ability to do 0-25% of their daily activities. Overall, these data suggest that experiencing mild AEs is common following vaccination with minimal impairment to daily activities.

### Females are more likely to report local AEs, regardless of age

For the influenza vaccine cohort, logistic regression models (**Figure 1A**) for probabilities of reporting any AE, any local, or any systemic AE, adjusted for sex, demonstrated that age was not significantly associated with AE reporting. Inclusion of an age-by-sex interaction term in the models (**Figure 1A**) revealed that the effect of age on the probability of reporting any AE, any local AE or any systemic AE after influenza vaccination of HCWs did not vary by sex. The average age of our HCW cohorts receiving the influenza vaccine (30.45 ± 6.9, range: 21-49) was relatively young and reproductive-aged (18-49).

**Figure 1.**
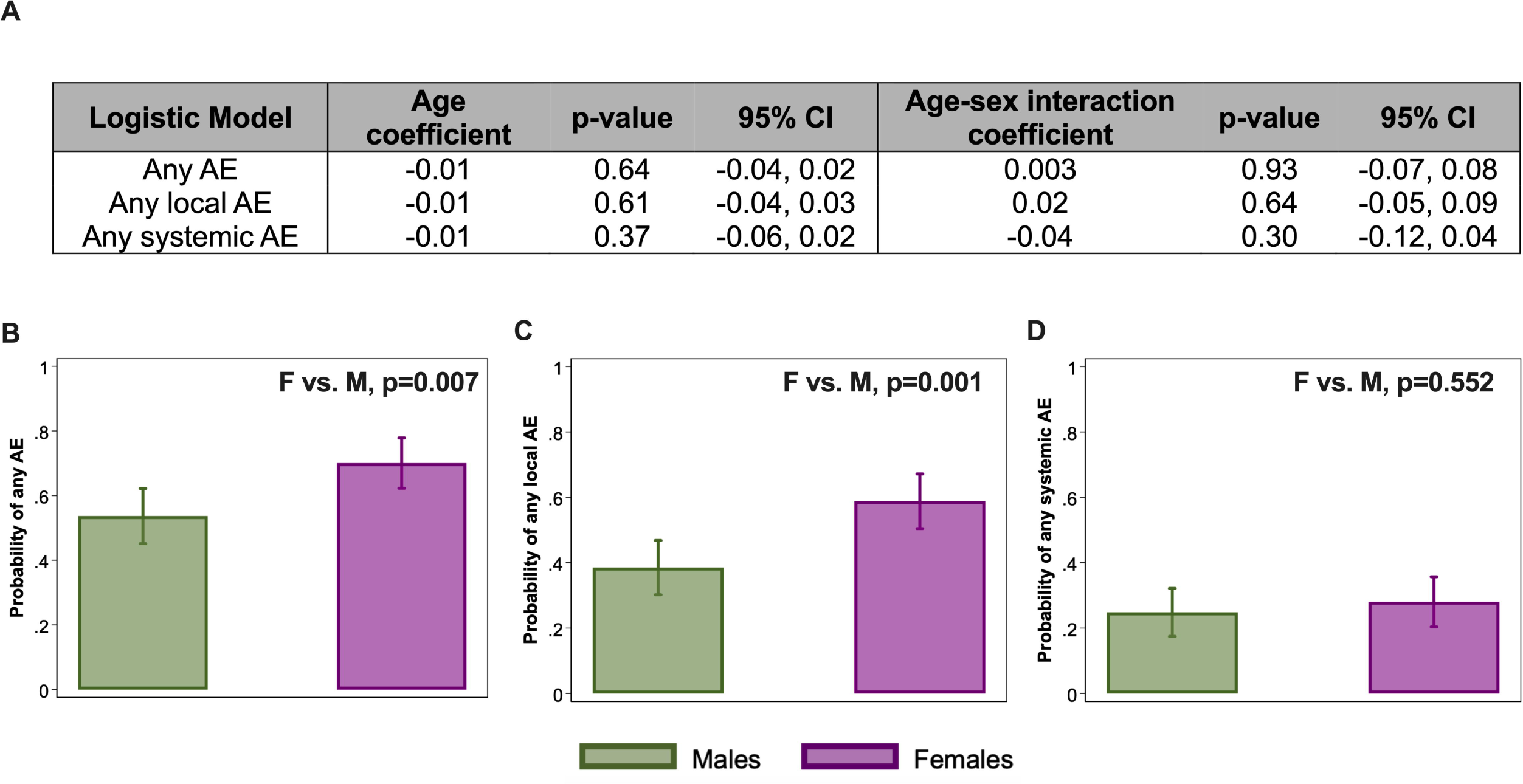
A greater proportion of female healthcare workers report local adverse events (AE) than males following annual inactivated quadrivalent influenza vaccination, regardless of age. A total of 300 quadrivalent influenzas vaccines were administered (151 males and 149 females) to healthcare workers (HCWs) from the 2019-2022 seasons, but AE data were only available for 265 of those participants (132 males and 133 females). (A) Logistic regression models for any AE, any local, and any systemic AE were used to assess the effect of continuous age, after adjusting for sex, or with an age-sex interaction term. Coefficients and p-values are shown for the models. Age and age-sex interactions were not significantly associated with the probability of reporting AEs following quadrivalent influenza vaccination; therefore, we focused on the effect of sex. (B-D) We performed sex-disaggregated analyses of local and systemic AE using logistic regression models to compare probabilities for (B) any AE, (C) any local AE, or (D) any systemic AE. Probabilities of AEs along with 95% confidence intervals are shown along with p-values for sex comparisons. P<0.05 was considered statistically significant.

Logistic regression models for probabilities of reporting any AE (at least one local or systemic AE) among influenza vaccine recipients across all three seasons showed that females had a significantly greater probability of reporting AEs compared to males (OR=2.02, 95% CI: 1.2-3.4, p=0.007; **Figure 1B**). Females had a significantly greater probability of reporting any local AE compared to males (OR=2.28, 95% CI: 1.4-3.7, p=0.001; **Figure 1C**), whereas the probability of reporting any systemic AE was comparable between males and females (OR=1.18, 95% CI: 0.68-2.0, p=0.552; **Figure 1D**).

For the COVID-19 vaccine cohort, logistic regression models (**Figure 2A**) also did not find a significant association of age with the probabilities of reporting any AE, any local, nor any systemic AE after adjusting for sex among the COVID-19 vaccine cohort. The probability of reporting any AE, any local AE, or any systemic AE was similar across ages for both male and female HCWs following bivalent COVID-19 vaccination (**Figure 2A**). The average age of our HCW cohort receiving the bivalent COVID-19 vaccine (38.4 ± 12.3; range: 22-75) was relatively young with less than 25% of the participants over 50 years-old. Taken together, these data suggest that age does not contribute to the probability of reporting an AE, regardless of vaccine type.

**Figure 2.**
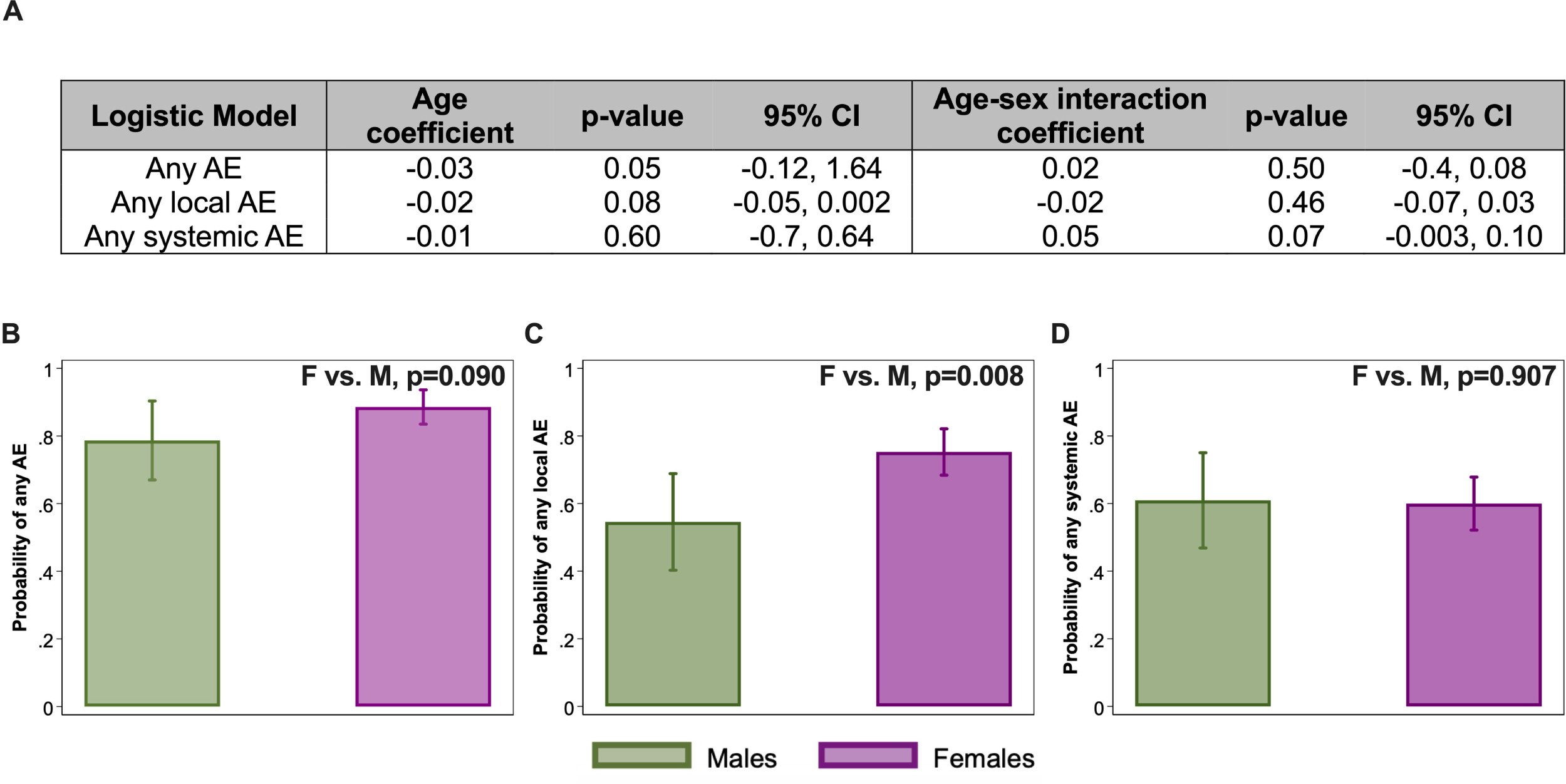
Females, regardless of age, have a higher probability of reporting any local adverse event (AE) than males following the bivalent Omicron ancestral/BA.5 COVID-19 vaccination. A total of 196 HCWs (46 males and 150 females) received bivalent COVID-19 vaccines, enrolled, and completed AE data in the 2022-23 season. (**A**) Logistic regression models for any AE, any local, and any systemic AE were used to assess the effect of continuous age, after adjusting for sex, or with an age-sex interaction term. Coefficients and p-values are shown for the models. Age and age-sex interactions were not significantly associated with the probability of reporting AEs following bivalent COVID-19 vaccination; therefore, we focused on the effect of sex. (**B-D**) We performed sex-disaggregated analyses of local and systemic AE using logistic regression models to compare probabilities for (**B**) any AE, (**C**) any local AE, or (**D**) any systemic AE. Probabilities of AEs along with 95% confidence intervals are shown along with p-values for sex comparisons. P<0.05 was considered statistically significant.

There was no significant difference in the probability of reporting any AE following bivalent COVID-19 vaccination between males and females (OR=2.14, 95% CI: 0.89-5.1, p=0.09; **Figure 2B**). Female HCWs, however, had a significantly greater probability of reporting any local AE compared to males (OR=2.57, 95% CI: 1.3-5.2, p=0.008; **Figure 2C**). Systemic AEs were similarly reported by males and females (OR=0.96, 95% CI: 0.49-1.9, p=0.907; **Figure 2D**).

### Sex differences of AEs are consistent across race categories in response to the influenza vaccination

Among the influenza vaccine HCW cohort, there were more females than males among those identifying as White (n=84, 52.5% females; n=76, 47.5% males) or Black (n=21, 61.8% females; n=13, 38.2% males; **Figure 3A**). For those identifying as Asian (n=22, 43.1% females; n=29, 56.9% males) or other (n=5, 27.8% females; n=13, 72.2% males), there were more males than females (**Figure 3A**). The logistic regression model for the probability of any AE with an interaction term for race and sex, adjusted for age, revealed that regardless of race, females consistently had greater probabilities of reporting any AE compared to males (**Figure 3B**). The interaction model did not show statically significant differences for reporting of any AE between males and females across race categories except for Black/African Americans, likely due to smaller sample sizes (**Figure 3A-B**). The probability of reporting any local AE consistently had a female bias with White and Black/African American females having significantly greater probabilities of reporting any local AE (**Figure 3C**). Systemic AEs were not significantly different between males and females across all race categories (**Figure 3D**). Race-disaggregated analyses were not performed with the COVID-19 AE dataset due to insufficient numbers of males to compare against females across race/ethnicity categories in the cohort.

**Figure 3.**
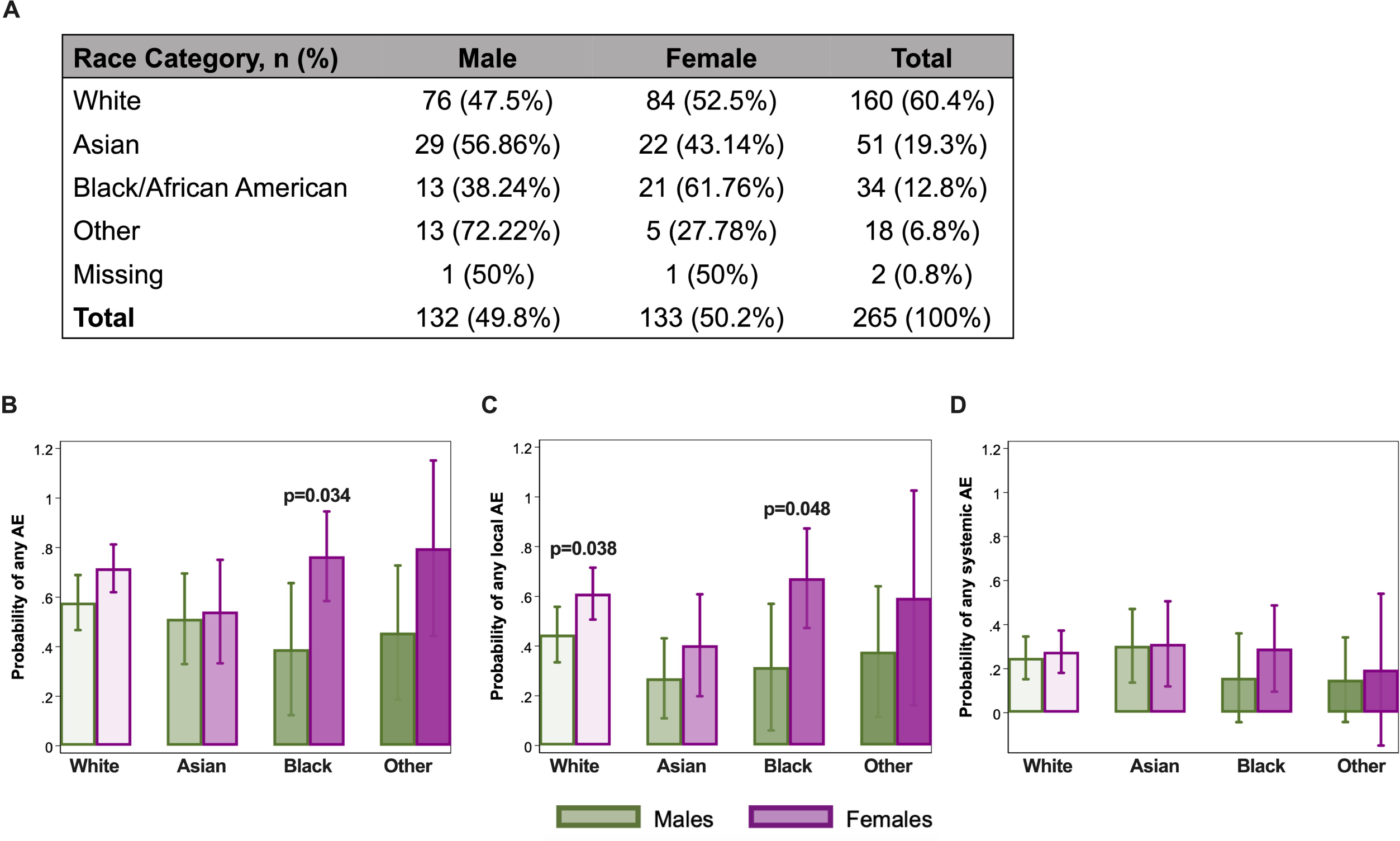
Females have a higher probability of reporting local adverse events (AEs) than males following annual inactivated quadrivalent influenza vaccination, regardless of race. (**A**) Descriptive table showing the breakdown of race categories by biological sex across the cumulative seasonal influenza seasons among healthcare workers. (**B**) Age-adjusted logistic regression model with a race and sex interaction term for any AE, (**C**) any local AE, or (**D**) any systemic AE. Probabilities of AEs along with 95% confidence intervals are shown along with p-values for sex comparisons. P<0.05 was considered statistically significant.

### Hormonal birth control use among females was not associated with the probability of reporting AEs

Birth control use (e.g., barrier method, oral contraceptives, IUD, etc.) data was collected at enrollment for 132 females with 55% (n=72) on birth control and 45% (n=60) not on birth control (**Figure 4A**) in the influenza vaccine cohort only. The average ages of female birth control users and non-users were 32.5 and 30.3 years, respectively. Among birth control users, hormonal birth control was the most common method with 44% (n=32) using oral contraceptives and 36% (n=26) using IUDs. Females who used the barrier method (n=2 of 72) were excluded to limit the birth control users to those on hormonal methods. Using logistic regression models, we assessed if the probability of reporting any AE (**Figure 4B**), any local (**Figure 4C**), and any systemic AE (**Figure 4D**) differed by hormonal birth control use among females. The probabilities of reporting any AE (OR=1.43, 95% CI: 0.66-3.1, p=0.36), any local (OR=0.94, 95% CI: 0.46-1.9, p=0.85), or any systemic AE (OR=1.78, 95% CI: 0.8-3.95, p=0.16) were similar between females using and not using birth control. These data suggest that exogenous hormones are no more likely than endogenous hormones to impact experiencing AEs in young adults of reproductive ages.

**Figure 4.**
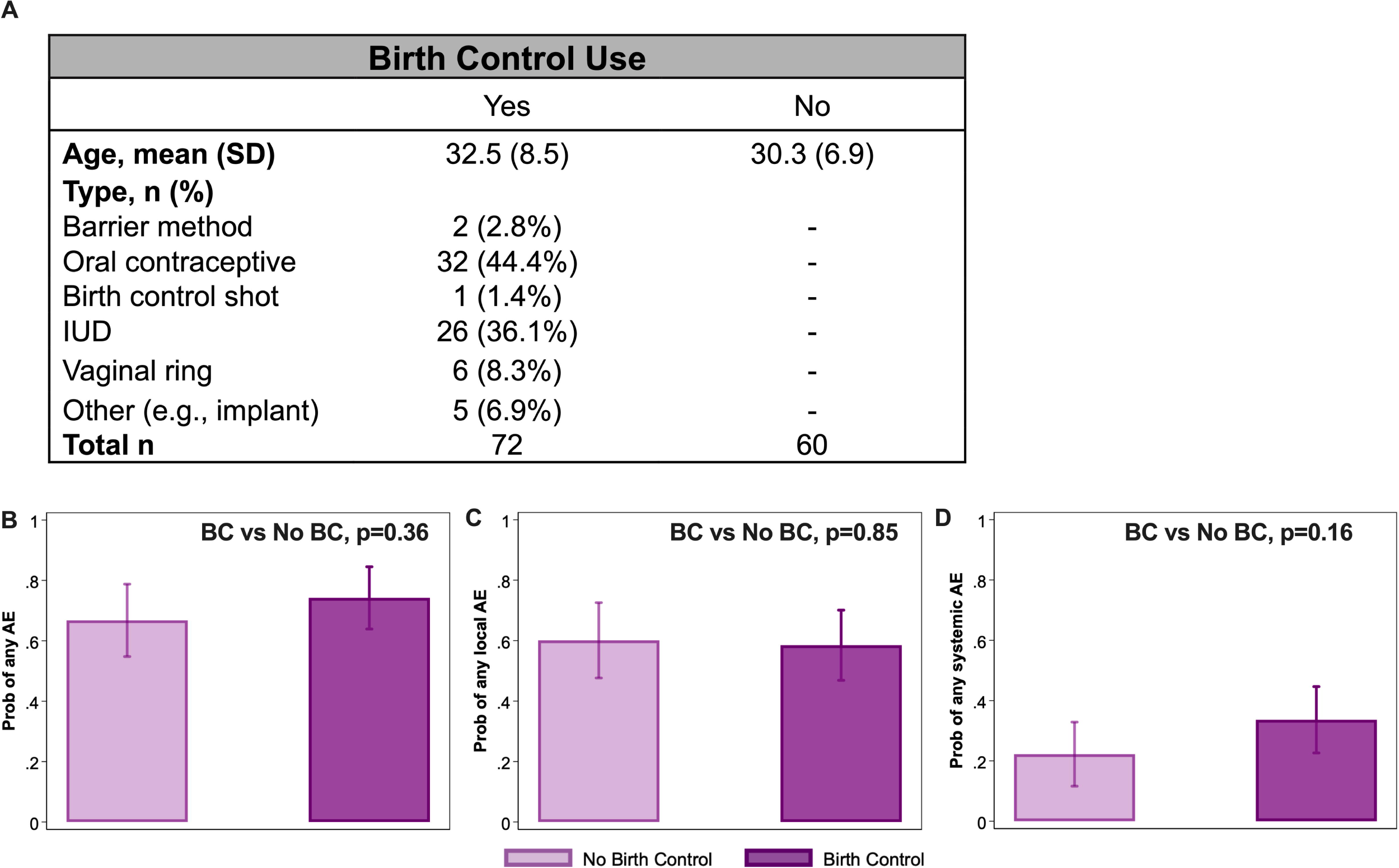
Hormonal birth control (BC) use among female healthcare workers did not impact the probability of reporting adverse events (AEs) after influenza vaccination. (**A**) Table of female healthcare workers, disaggregated by hormonal birth control use. Logistic regression models were used to examine probabilities of local and systemic adverse events (AE) following (**B-D**) annual influenza vaccination from 2019-2022 seasons among healthcare workers. Comparisons of probabilities for (**B**) any, (**C**) any local, or (**D**) any systemic AE following influenza vaccination are shown, respectively, with p<0.05 for birth control differences considered statistically significant. Probabilities of AEs along with 95% confidence intervals are shown.

### Women are more likely to report daily life disruptions following COVID-19 vaccination

Analyses of the open-ended survey answers completed by n=195 bivalent COVID-19 vaccine recipients revealed that female HCWs were more likely to report disruptions in their daily activities than males after receipt of the COVID-19 vaccination. There were 58 (38.7%) women who mentioned experiencing sleep disruption or changes in daily routine due to AEs following vaccination as compared to 14 (31.1%) men. Women also mentioned that AEs affected their ability to take care of their families.

> *[The vaccine] made me sleep for 10 hours, with other symptoms, usually sleep 7-8 hr. Felt harder to do activities of daily living and needed to lie down the next afternoon [White_W_4]*
>
> *Didn’t clean up from dinner or do my usual heavy lifting in terms of getting the kids to bed. [White_W_31]*
>
> *[I was] not able to take care of my baby. [White_W_113]*

### Women and men differed in how they responded to AEs

Women and men differed in how they responded to AEs. There were 36 (24%) women who reported self-administration of medications to mitigate symptoms of their AEs after receiving the COVID-19 vaccine as compared to 7 (15.6%) men, which is likely a reflection of more women experiencing AEs overall. Ibuprofen, acetaminophen, and over-the-counter pain relief medications were commonly used among those who did self-treat.

> *I took the recommended dose (2 capsules) of Tylenol every 6 hours for 18 hours starting 24 hours after the vaccination. [White_W_69]*
>
> *[I took] Acetaminophen and ibuprofen as well as increased electrolytes and hydration. [White_W_48]*

Some women expected to experience AEs and intentionally scheduled their COVID-19 vaccinations prior to days off; for example, scheduling the vaccine on a Friday so they did not have to miss work should they experience an AE.

> *Just know to plan for a Friday. Glad it was the weekend as I would have missed a day of work. I got the shot on a Friday on purpose as I had a bad reaction before with one of the others. [White_W_71]*
>
> *Planned the timing of the injection based on previous reactions so that I would be able to rest at home, [White_W_57]*

## DISCUSSION

We performed sex- and gender-disaggregated analyses of AE survey data for two different vaccines—the quadrivalent influenza and bivalent COVID-19 vaccine—to examine vaccine outcomes and vaccine-related behaviors among a cohort of adult HCWs, which can inform public and occupational health vaccine strategies and policies. Our study supports the existing evidence that influenza and COVID-19 vaccines do not cause serious AEs with localized, mild AEs being the most common experience[38, 39]. The bivalent COVID-19 vaccine recipients reported higher rates of AEs compared to influenza vaccine recipients in our cohorts, consistent with a retrospective analysis of VAERS data[40]. Increased AE reporting rates among COVID-19 vaccine recipients may be potentially confounded by the heightened scrutiny and vaccine hesitancy against mRNA COVID-19 vaccines at the time but is nonetheless important to note for public health and education purposes. While the term “adverse event” may suggest harmful or negative effects, non-serious AEs are normal and healthy manifestations of the immune system’s response to the vaccine antigen[11]. Transparent and consistent reporting of AEs is imperative to normalize these vaccine-related experiences, mitigate fear and misinformation, and encourage vaccine uptake.

Studies identifying age effects on the reporting of AEs are most common among older aged vaccinees. For example, Shapiro et al. found a female-specific effect where the probability of reporting any AE, either local or systemic, significantly decreased with increasing age for females, but not for males, 6-8 days after influenza vaccination among older adults (75+)[2]. Xiong et al. found that the proportion of COVID-19 vaccine AEs was greatest among younger adults (18-64) while the proportion of serious AEs was greatest among older adults[41]. Our analyses did not identify a significant age effect on the reporting of AEs following influenza or COVID-19 bivalent vaccination, likely because the cohort was predominately younger and reproductive-aged individuals.

Sex-disaggregated analyses revealed that female HCWs were significantly more likely to report local AEs, but not systemic AEs, after receipt of either the influenza vaccine or COVID-19 vaccine. Similarly, an active surveillance study of predominately younger adults (20-49) in South Korea reported females having significantly more AEs, local or systemic, after receiving the first dose of ChAdOx1 nCoV-19(AstraZeneca/Oxford) vaccine as compared to males based on self-reported survey results at 3 days post-vaccination[42]. In another highly vaccinated population of older adults (75+), females had greater probabilities of reporting local AEs, but not systemic AE, compared to males 7 days after receipt of the high-dose quadrivalent influenza vaccine as measured by AE surveys[2]. Xiong et al. utilized real-world data extracted from the Vaccine Adverse Event Reporting System (VAERS) to reveal that although more adult (18-64) females reported AEs within one week of COVID-19 vaccination, males had 1.5 times greater odds of reporting serious AEs[41].

Further disaggregation by sex and self-reported race demonstrated that females, regardless of race, consistently had a higher probability of experiencing local AEs with sex comparisons of White or Black participants reaching statistical significance. The probability of experiencing a systemic AE was comparable between sexes, regardless of race. While we did not find differences between racial categories, consideration for race and ethnicity analyses are important for vaccine studies. Race is not a biological variable associated with AEs, but race and ethnicity have been widely reported as important predictors of vaccine behaviors and perceptions[1, 43-47]. In a survey study of over 10,000 HCWs, Momplaisir et al. found that COVID-19 vaccine hesitancy was highest among Black and Hispanic or Latino HCWs when compared to White HCWs with worries about side effects as the most frequently cited reason[43].

These sex differences in adverse events are not specific to vaccines and have also been reported for other therapies, such as cancer immunotherapies, suggesting an underlying biological mechanism[48-50]. In a study of small-cell lung cancer patients receiving chemotherapy, Singh et al. reported that although a greater proportion of females were found to have more chemotherapy toxicity (e.g., hematologic toxicity, stomatitis, and vomiting) than males, females also had higher response rates and longer median survival time[49]. Unger et al. performed a meta-analysis of 202 clinical trials of cytotoxic therapy, immunotherapy, and targeted therapies[42] and found that females had significantly greater odds of severe toxicity and had a 66% increased risk of symptomatic AEs compared to males. Unlike vaccines that are mass-produced, personalized medicine may provide new avenues for other therapies or drugs, especially those with more severe AEs, to address sex differences in AEs[51, 52].

The role of sex steroid hormones in the manifestation of vaccine AEs for males and females is not clearly understood. Although our study was not designed to evaluate the hormonal and immunological responses associated with post-vaccination AEs, we were able to use hormonal birth control data (e.g., contraceptive use, IUD, implant, etc.) among females to assess if exogenous hormones mediated the reporting of AEs. Our data revealed that reporting of AEs did not differ by hormonal birth control use among young, reproductive-aged female HCWs. This may be due to reproductive-aged females already having sufficient endogenous sex steroid hormones such that birth control (i.e., exogenous hormones) did not change experience of vaccine AEs. Whether exogenous hormone use among postmenopausal women affects experiences of vaccine AEs requires consideration.

While more studies are implementing sex-disaggregated analyses, gender-disaggregated analyses are sparse in biomedical research due to the lack of an objective, standardized methodology for measuring gender and persistent misunderstanding of gender and sex. Examining vaccine outcomes and behaviors with a gender lens can inform public health messaging strategies and improve vaccine uptake. For instance, studies have found women have greater influenza and COVID-19 vaccine hesitancy compared to men worldwide[26, 27, 44, 53-55]. A survey study of HCWs in New York found that men had a higher likelihood of planning to get the COVID-19 vaccine within the next six months than women[31]. Although pregnancy and breastfeeding have been hypothesized as factors contributing to reduced vaccine uptake among women, Ciardi et al. did not find differences in vaccination uptake between reproductive-aged and non-reproductive-aged women[31]. HCWs are a unique population with increased access to accurate vaccine and medical information, yet vaccine hesitancy, particularly due to AEs, is still persistent even when vaccines are mandatory because of occupational exposure and spread[29, 30, 32].

Gender differences in vaccine behaviors pertaining to AEs are understudied. To our knowledge, we are among the first to utilize both quantitative and qualitative measures through open-ended survey questions to provide insight and context for the findings in a vaccine AE study. Our qualitative thematic analysis of open-ended answers revealed that women were more likely to seek out self-treatment (e.g., over-the-counter pain medications, ibuprofen, and acetaminophen) for their AEs and to experience disruptions in their daily routines than men after COVID-19 vaccination. In the meta-analysis conducted by Beyer et al.[9], the authors reported more women experiencing moderate to severe levels of inconvenience after influenza vaccination. In our study, experience of AEs from prior vaccinations motivated some women, but not men, to schedule their COVID-19 vaccinations on a day prior to their scheduled time off. Further interrogation of these gender differences in vaccine AE-related behaviors may inform vaccine campaign strategies or messaging, particularly among working-aged populations.

According to the 2019 U.S. Census Bureau women comprised 76% of healthcare jobs with 85% of nursing and health aide positions held by women[56]. We found that women HCWs were more likely to experience AEs than men and were more likely to seek out self-treatment and/or schedule vaccination prior to their days off from work. In a California survey of over 2,000 HCWs studying COVID-19 vaccine side effects, 28% experienced side effects that were disruptive to work and 18% missed work[57]. The authors also found that 6.7% of physicians missed work as compared to 21.2% of other HCW roles. Presenteeism, working despite feeling unwell or sick, and absenteeism are linked to occupational expectations and pressures that may differ across HCW roles, and can impact the quality of patient care, occupational burnout, and employee morale[57]. With nearly 9 million HCWs nationwide receiving mandated vaccinations, we can expect that millions of workers (the majority of whom are women) will experience AEs annually with potential occupational health and labor force implications, including increased vaccine hesitancy, missed work, and disruptions to recognized time off. Our data add gender to the list of factors that need to be considered in policies surrounding mandatory vaccines, including, for example, receipt of paid medical leave.

## LIMITATIONS

There are several limitations to this study. First, the enrollment criteria (e.g., age) were different and did not allow for direct comparison between the two cohorts. The sample size of males and females enrolled were only pre-specified and balanced for the influenza vaccine cohort and not the COVID-19 vaccine cohort; therefore, the COVID-19 vaccine cohort may be more representative of the HCW demographics at JHHS. Highly vaccinated HCWs are more likely to be biased towards vaccine acceptance and the interpretations made from this unique demographic may not be applicable to non-HCW populations. Second, the criteria and definitions for local and systemic AEs used may differ from other studies. AEs were surveyed two days post-vaccination, so we were unable to assess AEs after administration of the questionnaires. Lastly, biological samples were not collected from participants; therefore, we were unable to study the immunological mechanisms by which sex causes differences in AEs.

## CONCLUSIONS

Our AE survey study of HCWs following influenza or COVID-19 vaccination demonstrates that females were more likely to experience local AEs than males. Women were more likely to experience interruptions in their daily routines and to self-treat AEs. Additionally, more women reported scheduling their vaccines on a day before their scheduled time off in anticipation of AEs. These data highlight the importance of considering sex and gender in public health and occupational health vaccine strategies and communications, particularly when targeting the predominately female healthcare workforce. Further sex- and gender-disaggregated research is needed to build more equitable and effective vaccine strategies with consideration for differences in AEs. Development of such strategies is not only important for seasonal vaccination planning, but also for planning effective vaccination campaigns for HCWs during pandemics.

## ARTICLE INFORMATION

## Data Availability

All data produced in the present study are available upon reasonable request to the authors.

## Acknowledgements

We kindly thank the healthcare workers of the Johns Hopkins Health System for their participation in our studies and the clinical coordinators who helped with recruitment and data collection.

## Contributors

AY, PS, HK, JRS, KZJF, RER, AP, SLK, and RM contributed to the conception and design of the study. Data collection and statistical analyses were done by AY, NW, HK, and KZJF. AY, SLK, and RM wrote the first draft of the manuscript. All authors contributed to the critical review, interpretation of the results, and revision process.

## Patient and public involvement

Patients and/or the public were not involved in the design, or conduct, or reporting, or dissemination plans of this research.

## Patient consent for publication

Not required.

## Ethics approval

Our study involves two cohorts of human participants, which were approved by the Johns Hopkins Institutional Review Boards (IRB00259171, IRB00091667). Consent was obtained for all participants as part of the enrollment process.

## Provenance and peer review

Not commissioned; externally reviewed.

## Data availability statement

Data may be obtained from a third party and are not publicly available.

## Supplemental material

This content does not have supplemental materials.

